# Risk factors for anaemia among pregnant women: A cross-sectional study in Upper East Region, Ghana

**DOI:** 10.1101/2024.03.21.24304692

**Authors:** Clotilda Asobuno, Silas Adjei-Gyamfi, Felix Gumaayiri Aabebe, John Hammond, Chansathit Taikeophithoun, Norbert Ndaah Amuna, Tsunoneri Aoki, Hirotsugu Aiga

## Abstract

**Background:** Anaemia in pregnancy (AIP) is a public health concern due to its devastating effects on women and their unborn babies, resulting in increased maternal and neonatal deaths in developing countries. Despite several Ghanaian health policies to combat AIP, AIP is still on the rise. It becomes imperative to identify geographic-specific factors for developing appropriate interventions for the management of AIP. However, Kassena Nankana West District (KNWD) in the Upper East Region of Ghana lacks a study on anaemia risk factors, therefore, this study estimated the prevalence and risk factors for anaemia among pregnant women in the district.

**Methods:** A household-based cross-sectional study was conducted on 376 pregnant women in their third trimester from February to March 2023. Anthropometric, obstetric, sociodemographic, and health facility resource characteristics were collected using structured questionnaires and antenatal records. Mixed-effect logistic regression was used to identify independent factors of anaemia at 95% confidence interval.

**Results:** Prevalence of AIP was 53.9%. Mild, moderate, and severe anaemia prevalence was 16.9%, 35.3%, and 1.7% respectively. Malaria infection during pregnancy (aOR=1.563; 95%CI:1.087 – 2.475) and accessing health facilities without trained laboratory personnel (aOR=5.271; 95%CI:1.641 – 16.93) were associated with increased odds of AIP. Belonging to the major ethnic group (aOR=0.431; 95%CI:0.280 – 0.675), accessing health facilities without laboratory services (aOR=0.151; 95%CI:0.047 – 0.487), and accessing health facilities without sulphadoxine-pyrimethamine drugs (aOR=0.234; 95%CI:0.061 – 0.897) in KNWD were also associated with decreased odds of AIP.

**Conclusion:** AIP prevalence remains high in the KNWD. Maternal and health facility-related factors were responsible for anaemia in the district. These factors are preventable. Therefore, health facility strengthening and enhanced strategies for malaria prevention are recommended for anaemia control in the district.

## Introduction

Anaemia affects all populations in the world, especially pregnant women, and women in their reproductive age (1). It is one of the most severe and prevalent nutritional deficiencies of public health concern among pregnant women in low and middle-income countries (LMICs) (2). The World Health Organization (WHO) describes anaemia in pregnancy (AIP) as decreased haemoglobin (Hb) level of <11g/dL and is further classified into severe, moderate, and mild (3). Globally, 50,000 young women die each year in pregnancy and childbirth due to anaemia (4). In sub-Saharan Africa and Southern Asia, anaemia contributes to about 20% of all maternal and perinatal mortalities (3). Additionally, anaemia does not only reduce work productivity during pregnancy but also leads to adverse pregnancy outcomes such as low birth weight, premature delivery, stillbirths, postpartum haemorrhage, and postpartum morbidities (3,5–7).

Globally, anaemia is disproportionately distributed and affects about 40% to 60% of all pregnant women in LMICs (5). This high prevalence of LMICS is associated with iron deficiency thereby causing more than 50% of pregnancy-related anaemia cases and affecting nearly two billion population across the globe (3,8). A systematic review analysis in some LMICs including Burundi, Togo, and India reported a composited 50.3% prevalence of anaemia. The same study projected that, by 2025 anaemia prevalence among pregnant women in those countries (Burundi, Togo, and India) would reach 66.8%, 60.4%, and 60.3% respectively (9). In Ghana, the Demographic and Health Survey in 2014 estimated anaemia among pregnant women in the country at 45% (10) while a retrospective study in northern part of the country reported anaemia prevalence of 50.8% (11). Data from the Kassena-Nankana West District Health Directorate indicates that from 2018 to 2021 the proportion of registrants with anaemia during antenatal care had significantly increased from 33% to 46%. Moreover, the trend prevalence of gestational anaemia at the third trimester of pregnancy in the same district was 31%, 46.5%, 46.3%, and 50.9% from 2018 to 2021 (12). This highlights the need to conduct studies to identify factors associated with anaemia in the district.

Several policies and strategies on anaemia prevention have been adopted by many developing countries in the world. The global strategy for anaemia reduction included vector elimination via the distribution of insecticide-treated bed nets, provision of chemoprophylaxis treatment, and periodic treatment with anthelminthic for all women of childbearing age including pregnant women in endemic areas (1,3,8,11). Another strategy is girls’ iron folic acid tablet supplementation to prevent anaemia, especially in areas with increasing adolescent deliveries and early marriages (4). Iron and folic acid supplementation for pregnant and postpartum women have been evidenced to yield optimal results in the prevention of anaemia (3,4). Additionally, Scaling Up Nutrition (SUN) Movement (Strategy 3.0) has created a positive impact on nutrition through education, water, hygiene practices, and environmental sanitation, as well as strengthening food, health, and social protection systems in most developing countries (13). Ghana has utilized the SUN concept in community-based health planning and services (CHPS) implementation to reach out to all populations. This initiative brought health services to the doorsteps of the people for health workers to identify and manage minor illnesses including anaemia among pregnant women in Ghana. Despite these policy efforts, anaemia is still on the rise (11,14).

The contributing factors of anaemia in pregnancy are largely classified into individual, cultural, nutritional, socioeconomic, and infection-related factors (5,8). Some reported studies in LMICs regarded human immunodeficiency virus infection, primigravida, multiparity, and teenage pregnancies as prominent determinants of anaemia during pregnancy (5,11,15–18). In Africa and Ghana, some predictors of anaemia during gestation are elderly pregnant women, low household wealth index, and illiteracy (no formal education) (5,19,20). In the Kassena-Nankana West District (KNWD), relevant scientific studies on risk factors of pregnancy-related anaemia for informing appropriate interventions are lacking. It is not clear whether there are some service delivery factors, inherent maternal factors, and lack of client adherence to medications that are leading to the persistent rise in anaemia among these pregnant women. Notwithstanding, most studies in Ghana do not consider health facility-related determinants of anaemia for making informed decisions (11,14,20). It becomes more prudent to dig into health facility-based and maternal determinants that could be associated with anaemia during the period of pregnancy. This study therefore seeks to estimate the prevalence and risk factors of anaemia among pregnant women in the KNWD, Upper East Region (UER) of Ghana.

## Materials and Methods

### Study design

This study was a household cross-sectional study conducted to estimate anaemia prevalence and identify the risk factors for anaemia among pregnant women in KNWD of UER, Ghana.

### Study area

The study was conducted at KNWD in the UER of Ghana which was carved out of Kassena-Nankana Municipality in 2007 and launched on 29^th^ February 2008. This district’s 2021 population projected from the National Population and Housing Census was 89,043 with a yearly growth rate of 2.4% (21). KNWD shares a border with Bulsa District in the southwest, Sisala District in the west, and Burkina Faso in the north, Bongo and Bolgatanga Districts in the east and northeast respectively. There are two main climatic seasons found in the district; a dry season and a rainy season with average rainfall between 850 – 1000mm. Kassenas and the Nankanas are the two prominent ethnic groups. The major income source in the district is farming and petty trading. There are nine health centers, one district hospital as a referral center, 51 CHPS compounds, and 3 private health facilities (12). These facilities provide health services to all 115 communities in the district (21). The total antenatal care (ANC) visits in 2021 was 17,566. The total number of women in their reproductive age is estimated at 21,370 (24%), with yearly estimated pregnancies of 3,562 (4%) (12).

### Study population

This study interviewed 395 pregnant women who were registered and attending antenatal care in the district. Pregnant women having maternal and child health record books (MCHRBs) and were in their third trimester, were included in the study. Those with haematological problems or who were seriously sick during data collection were excluded from the study.

### Sample size and sampling methods

The sample size (n) was calculated using the Cochran formula (22); 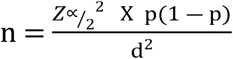

The study employed the prevalence (p) of AIP in a neighbouring district (Kassena Nankana East Municipal) in the UER of Ghana estimated at 42.7% (14), margin error (d) of 5%, and standard normal variate (*Z*_∝/2_) at 95% confidence level of 1.96, the sample size was initially estimated at 376. Due to incomplete responses, 5% of the sample size was added (7) to produce an approximated final sample size of 395 (= 376/ [1 – 0.05]).

Of the 64 health facilities in the KNWD, only 10 health facilities provide daily ANC services in the district. During sampling, these 10 major health facilities were selected for the study. Probability proportional to size technique was used to determine the sample sizes for each health facility. Each facility’s ANC register was obtained to line-list all pregnant women who were eligible to form a sampling frame. Thereafter, a simple random sampling method was used to select the respondents from each health facility. The addresses (including contact numbers) of the recruited respondents were then taken to trace these respondents to their homes.

### Data collection

A pretested structured questionnaire was used to collect the data from the selected health facilities and pregnant women in the comfort of their homes in February and March 2023. The questionnaire was designed using Kobo collect. Data was first collected on health facility’s resources for anaemia measurement and prevention including laboratory services on blood testing, and availability of iron and folic acid (IFA) supplements, sulphadoxine-pyrimethamine (SP) drugs, and MCHRBs. The next phase of the data collection was at the households of respondents. Before household data collection commenced, the trained research assistants obtained the addresses and contacts (telephone numbers) of respondents and/or representatives from the ANC register. The midwives in each facility helped the research assistants to obtain this information. The research assistants then visited the homes of the recruited respondents after booking an appointment with them. Data on Hb levels, antenatal, and anthropometry were confirmed from the MCHRBs, and other information including sociodemographic, obstetric, and socioeconomic characteristics were collected through structured interviews.

### Study variables

The dependent variable for this study was AIP in the third trimester classified as; (i) Anaemia (Hb: < 11 g/dl) and (ii) No anaemia (Hb: ≥ 11 g/dl). Anaemia status was further classified into mild (Hb: 10.0 –10.9g/dl), moderate (Hb: 7 – 9.9 g/dl), and severe anaemia (Hb: < 7 g/dl) (3). The sociodemographic, socioeconomic, anthropometric, antenatal, obstetric, and availability of resources for anaemia measurement and prevention at the health facilities were the independent variables. Most of the independent variables including iron-folic acid intake, anthelminthic drugs intake, SP doses intake, human immunodeficiency virus (HIV) infection, malaria during pregnancy, and availability of health facility resources (SP, iron folic acid, anthelmintics, antimalaria drugs, functional laboratory machines, and trained laboratory personnel) were grouped as Yes/No. Pre-pregnancy (first-trimester) BMI was classified into underweight (< 18.5 kg/m^2^), normal (≥ 18.5 to 24.9 kg/m^2^), and overweight/obese (≥ 25.0 kg/m^2^) using the WHO classification (23). Gravida was grouped into primigravida (0–1 pregnancy) and multigravida (≥ 2 pregnancies). Parity was classified into primipara (0–1 delivery) and multipara (≥ 2 deliveries) (6). Based on maternal gestational age, ANC initiation was summarized into first (≥13 weeks), second (14 – 27 weeks), and third (28 – 41 weeks) trimesters (23). By employing principal component analysis, the wealth index of respondents was assessed using household properties and housing quality which was used as a proxy indication for socioeconomic status of the respondents. The wealth index was then categorized into five quintiles namely: poorest, poorer, middle, richer, and richest (6). Other variables such as ethnic group, religion, educational level, occupation, marital status, and household size were also summarized and categorized based on previous studies (1,6,7)

### Statistical analysis

Statistical analysis was conducted using STATA version 17.0 (Stata Corporation, Texas, USA) with all analyses determined at 95% confidence level. Chi-square or Fischer’s exact test was used to determine the bivariate association between anaemia status and each independent variable. In the multivariate analyses, mixed-effect logistic regression was used to identify risk factors for AIP by applying a simultaneous variables entry. The study used mixed-effect logistic regression because majority of the independent variables were nested at the health facilities level. All independent variables with significant bivariate association with anaemia (p < 0.05) were chosen as possible predictor variables for logistics analyses. Multicollinearity was checked among the predictor variables before employing them in the multivariate analyses. Variance Inflation factor and Spearman’s correlation coefficient were used during multicollinearity testing.

### Ethical considerations

The study obtained ethical clearance from the Nagasaki University School of Tropical Medicine and Global Health (TMGH) Institutional Review Board (approval number:NU_TMGH_2022_225_1), and the Ghana Health Service Ethics Review Committee (approval number: GHS-ERC:023/12/22). Permission was obtained from the UER Health Directorate, KNWD Health Directorate, and Heads of the health facilities before the data was collected. Written informed consent was obtained from all respondents and/or their parents/guardians after they were given information on the purpose of the study and what was required of them as study participants.

## Results

A total of 395 respondents were interviewed. However, 19 incomplete questionnaires were found during data cleaning. Hence, 376 respondents were used for the data analysis.

### Socio-demographic, antenatal, and obstetric characteristics of study participants

The ages of the pregnant women ranged from 16 to 50 years, with a mean age of 27 years (sd=6.28). A greater number of the respondents were married (93%) and were Christians (85%). The respondents were predominantly from the Kassena ethnic group (55%), while slightly above half of them were unemployed (51%). About 10% of them had no formal education (Table 1). More than one-quarter of the women (27%) received IFA tablets from their health facilities. While 76% of the pregnant women slept under ITNs the night before the interview, one-quarter of them (25%) had taken at least four doses of SP during pregnancy. Slightly above one-third of the women (35%) experienced malaria during their pregnancy period (Table 2).

### Availability of health facilities’ resources for anaemia measurement and prevention during antenatal visits

Over half of the pregnant women made ANC visits to health facilities without functional laboratory services (54%), while those who attended health facilities without haemoglobin testing machines were nearly one-third (30%). A greater proportion of pregnant women received ANC services from health facilities without trained laboratory personnel. More than half of the pregnant women (62%) accessed the health facilities having no stock of MCHRBs. Furthermore, 79% of the respondents accessed ANC in the health facilities having no stock of IFA supplements. Few pregnant women (12%) did not receive SP drugs from facilities they visited for ANC services (Table 3).

### Prevalence of anaemia among study participants

The mean haemoglobin level was found to be 10.85 g/dl (sd=1.36). The prevalence of anaemia was 53.9% (95%CI:48.5% – 58.8%) which was classified into mild, moderate, and severe anaemia corresponding to 16.9%, 35.3%, and 1.7% respectively (Table 1).

### Association of anaemia with exposure variables

As shown in Tables 1, 2, and 3, ethnicity (p = 0.002), malaria during pregnancy (p = 0.006), accessing health facilities without laboratory services (p = 0.023), accessing health facilities without haemoglobin testing machines (p =0.021), accessing health facilities without trained laboratory personnel (p = 0.024), and accessing health facilities without SP drugs (p =0.040) were associated with anaemia at the bivariate level.

### Associated risk factors for anaemia in pregnancy

After chronicling no multicollinearity issues among the six predictor variables that shown significant bivariate association with anaemia status, five of them namely ethnicity, malaria during pregnancy, accessing health facilities without laboratory services, accessing health facilities without trained laboratory personnel, and accessing facilities without SP drugs remained statistically significant at the multivariate analysis level (Table 4).

The findings suggest that pregnant women who had malaria infection during pregnancy had 1.563 times increased risk of getting anaemia (aOR=1.563; 95%CI:1.087 – 2.475). Women belonging to Kassena ethnic group had a reduced risk of anaemia, indicating 56.9% ([1 – 0.431] x 100) reduction in anaemia (aOR=0.431; 95%CI:0.280 – 0.675). Additionally, pregnant women who accessed health facilities without laboratory services (aOR=0.151; 95%CI:0.047 – 0.487) and without stock of SP drugs to supply to pregnant women (aOR=0.234; 95%CI:0.061 – 0.897) had decreased odds for anaemia. Lastly, pregnant women who visited health facilities without trained laboratory personnel were more likely to be anaemic (aOR=5.271; 95%CI:1.641 – 16.93).

## Discussion

AIP is a global public health concern due to its consequences on the lives of women and unborn babies such as low birth weight, preterm births, stillbirths, and maternal deaths (6,7,24,25). The World Health Assembly aims to reduce anaemia by 50% in 2025 (3), which motivates most countries to identify and address factors responsible for anaemia during pregnancy. Hence, this household-based cross-sectional study was conducted to estimate anaemia prevalence and the possible risk factors among pregnant women in KNWD.

This study’s mean haemoglobin level was 10.85g/dl with anaemia prevalence of 53.9%. Our study’s anaemia prevalence was higher than the global and East African prevalence of 41.8% (9,26) and the prevalence reported by the 2014 Ghana Demographic and Health Survey (42%), Sunyani municipality (40.8%), and Hohoe municipality (32.7%) of Southern Ghana (20,27,28). Furthermore, previous studies in Northern Ghana reported a similar prevalence to that of the present study (5,11) while a similar study in Northern Tanzania reported higher anaemia rate of 81.3% (29). Paired with this study’s results, diversities in the cultural settings, beliefs, and economic activities between the southern and northern parts of Ghana and across Africa might be the reason for variations in the anaemia prevalence. Also, the variations in anaemia status among participants could be explained by several factors including the type of ethnic group, malaria infection, and accessing health facilities without laboratory services, trained laboratory personnel, and SP drugs which were found to be statistically associated with anaemia during pregnancy in this study.

Women belonging to the major ethnic group offered protection against anaemia during pregnancy. Kassena serves as the major ethnic group in the KNWD. This study result is similar to other study findings in Israel (30) and Denmark (31,32). The Kassena ethnic group is mostly found in the capital town of the district, where most women practice petty trading and are economically better off compared to other ethnic groups in the KNWD (11). This might influence their food purchasing, food intake, and health-seeking behavior which could reduce the risk of anaemia among the Kassena pregnant women.

The study showed that women with malaria during pregnancy had an increased risk of anaemia. Similar to a study in Southern Ghana (28), malaria in pregnancy is associated with anaemia while some cross-sectional studies in Northern Ghana (5,14) are incongruent. Africa remains the breeding ground for malaria parasites especially in tropical areas like Ghana (28,33). In 2010, malaria contributed to the majority of all deaths and about 11% of maternal deaths in Ghana (33). Malaria during pregnancy affects the maternal and foetal tissues and breaks down the formation of maternal erythrocytes leading to anaemia (33,34) and consequently resulting in adverse pregnancy outcomes such as low birth weight and premature deliveries (6,7,35). Ghana adopted WHO-recommended strategies for malaria control such as the distribution of insecticide-treated bed nets and chemoprophylaxis treatment via the intake of SP drugs for the prevention of malaria through the free maternal care policy (20,36). However, commitment and adherence to these interventions in KNWD may be suboptimal which could predispose the women to malaria and risk them for anaemia during pregnancy.

Pregnant women who attended ANC services at health facilities without trained laboratory personnel were at higher risk of developing anaemia. The present study’s finding is parallel to a similar study in Tanzania (37). This finding could occur as untrained laboratory persons are more likely to wrongly read, interpret, and report laboratory results due to a lack of technical expertise which may risk pregnant women from receiving the appropriate interventions for possible anaemia detection.

Our study revealed that pregnant women who accessed ANC services at health facilities without laboratory services were less likely to develop anaemia. Laboratory services are essential for the early detection, diagnosis, and monitoring of anaemia in pregnancy. These services facilitate targeted interventions, such as iron supplementation and nutritional counseling, which are vital in preventing maternal and foetal complications associated with anaemia (20,38,39). Timely and appropriate use of laboratory tests significantly contributes to the overall better health outcomes of pregnant women and their unborn foetus. Despite the enormous importance of availability of laboratory services in curbing anaemia in pregnancy, majority of the health facilities in Kassena-Nankana West district do not have the basic laboratory equipment to conduct the services. Therefore, all pregnant women who attended health facilities without laboratory services are given referrals to the facilities with laboratory services to conduct haemoglobin and other laboratory tests. The laboratory service may be an indirect determinant of anaemia; however, it could depend on the ability of the pregnant women to adhere to the preventive and treatment protocol given to them following the test results that may determine the anaemia status of the women.

Lastly, women attending health facilities without SP drugs as unexpected were protected against anaemia during pregnancy which is inconsistent with a report from McClure and colleagues who asserted that facilities with higher stock of SP drugs reduced the risk of maternal anaemia (40). SP drugs are known for their efficacy in preventing and treating malaria during pregnancy, which serves as a significant risk factor for anaemia (33,36). So, a pregnant woman who religiously adheres to SP intake and other malaria preventive measures tends to protect against malaria and anaemia (20,36). Facilities without SP drugs may be an indirect contributor to anaemia in pregnancy, hence further studies should consider health facilities’ impact on anaemia by assessing the SP stock level.

### Limitations of the study

Sampling bias due to exclusion of pregnant women who lost or not having their MCHRBs, because the study collected some data directly from the MCHRBs. Additionally, pregnant women who were not in their third trimester were excluded from this study which could affect sampling amd anaemia prevalence. This study collected secondary data on haemoglobin levels, obstetric, and anthropometric characteristics documented in the MCHRBs as some of the data may be inaccurate due to mis-recording or mismeasurement leading to information bias. The study could not be generalized and may need a larger sample size to determine rare risk factors of anaemia. Lastly, since it is a cross-sectional study, it might not measure the seasonal findings of AIP.

## Conclusion

AIP was very high in the KNWD which were affected by maternal ethnic group, malaria during pregnancy, and health facilities-related factors. Enhancing malaria prevention and treatment measures is essential for addressing some of these factors in KNWD. Ghana Health Service should recruit more laboratory personnel and periodically conducts in-service training to update the staff on haemoglobin testing. Moreover, Ghana Health Service should provide health facilities in hard-to-reach communities and peripherals with well-equipped laboratories for testing haemoglobin and other laboratory tests to increase accessibility for minor ethnic groups and provide a conducive environment for anaemia treatment protocol. We recommend that future larger studies should be conducted to assess health facilities-related factors on anaemia during pregnancy.

## Data Availability

The data are available from the corresponding author upon reasonable request.

## Acknowledgements

The authors are very thankful to The Project for Human Resource Development Scholarship - Japan International Cooperation Agency (JICA); Nagasaki University School of Tropical Medicine and Global Health – Japan; and Kassena Nankana West District Health Directorate, Ghana Health Service – Ghana, for supporting this study. Our gratitude also goes to all data collectors, research respondents, and volunteers who greatly contributed to this study.

## Abbreviations

ANC: Antenatal care
AIP: Anaemia in pregnancy
BMI: Body mass index
CHPS: Community-based preventive services
Hb: Haemoglobin
HIV: Human immunodeficiency virus
IFA: Iron and folic acid
ITNs: Insecticide-treated bed nets
KNWD: Kassena Nankana West District
LMICs: Low-and middle-income countries
MCHRBs: Maternal and child health record books
SP: Sulphadoxine pyrimethamine
SUN: Scaling Up Nutrition
UER: Upper East Region

## Availability of data and materials

The data are available from the corresponding author upon reasonable request.

## Competing interests

The authors declared that they have no competing interests

## Funding

This study was supported by The Project for Human Resource Development Scholarship, Japan International Cooperation Agency (JICA), and Nagasaki University School of Tropical Medicine and Global Health, Japan.

## Authors’ contributions

CA, SAG, NNA, TA, and HA designed and conceptualized the study. CA, FGA, JH, and CT carried out the data collection as SAG, NNA, TA, and HA played supervisory roles. CA, SAG, FGA, JH, CT, and HA performed the data analysis and interpretation. CA, SAG, and HA drafted the initial manuscript. All authors reviewed and approved the final manuscript.

## Authors’ acronyms

Clotilda Asobuno (CA); Silas Adjei-Gyamfi (SAG); Felix Gumaayiri Aabebe (FGA); John Hammond (JH); Chansathit Taikeophithoun (CT); Norbert Ndaah Amuna (NNA); Tsunoneri Aoki (TA); Hirotsugu Aiga (HA).

## Supporting information

S1 Table 1. Socio-demographic characteristics of respondents and prevalence of anaemia

S2 Table 2. Antenatal and obstetrics characteristics of respondents

S3 Table 3. Availability of health facilities’ resources for anaemia measurement and prevention during antenatal visits

S4 Table 4. Mixed-effect logistic regression on the risk factors for anaemia in pregnancy

## Notes

### Competing Interest Statement

The authors have declared no competing interest.

